# Diagnostic Accuracy and Yield of Determine Urine TB LF-LAM® for Detecting Tuberculosis Among Adults and Children with Advanced HIV Disease in Nigeria

**DOI:** 10.1101/2025.09.13.25335679

**Authors:** Justin Onyebuchi Nwofe, Laura Ifunanya Madukaji, Daniel Chinenyeike Offie, Dickson Adeolu Adetoye, Emmanuel Adewale Ojo, Magnus Odido, Izuchukwu Ibeagha, Christian Okechukwu Ugwu, Daniel Oguh, Adeola Awolola, Olabamji Samuel Osho, Patrick Chike Okonkwo, Adebola Mary Onyenike, Josephine Okemutie Omo-Emmanuel, Isaac Onyebuchi Okohu, Chima Paschal Chima, Oyewale Oguntade, Olatunde James Kehinde, Anthony Ani, Amali Owoicho, Enenche Edigah, Ada Immaculata Chinweuba, Daniel Ogbuagu, Femi Emmanuel Owolagba, Eke Ofuche, Jay Osi Samuels

**Affiliations:** APIN Public Health Initiatives, Nigeria

**Keywords:** HIV, tuberculosis, diagnostic accuracy, LF-LAM, advanced HIV disease

## Abstract

**Introduction:** Tuberculosis (TB) diagnosis remains the weakest link in the care cascade, particularly among people living with HIV and AIDS (PLHIV) with advanced HIV disease (AHD), who are often cannot produce quality sputum for standard tests. Urine-based assays such as Determine Urine TB LF-LAM® offer a non-invasive, easy-to-collect alternative that can detect lipoarabinomannan antigen even in extrapulmonary TB, making them especially useful in severely immunocompromised individuals.

**Methods:** We conducted a cross-sectional diagnostic accuracy and yield of Determine Urine TB LF-LAM® against GeneXpert MTB/RIF as the reference standard among adults and children (<5 years) with AHD enrolled across 334 selected HIV treatment facilities in Nigeria between July 2022 and July 2025. We calculated the DY for LF-LAM among all AHD patients tested and, in the paired-testing cohort, determined its incremental diagnostic yield (IDY). Test proportions are presented with 95% CIs using the Wilson score method; discordance in paired results was assessed with McNemar’s test.

**Results:** Of the 17,155 AHD patients tested with LF-LAM on first urine samples, 5,212 were TB LF LAM positive, giving a diagnostic yield of 30.4%. Among 5,212 LF LAM positives, 2,074 (41%) of them were not presumptive through WHO four-symptom screening and would have been missed without LF-LAM. Among 3,138 patients tested with both GeneXpert and LF-LAM, GeneXpert detected 1,914 TB cases. The remaining 1224 GeneXpert-negative cases tested positive through TB LF-LAM, giving an incremental yield of 39% for LF-LAM over GeneXpert (1224/(1,914+1224); Wilson 95% CI) with significant discordance (McNemar p<0.001).

**Conclusion:** Determine Urine TB LF-LAM® substantially improved TB case detection among PLHIV with AHD in routine Nigerian HIV programs, including pediatric and severely immunocompromised patients who are unable to produce sputum or with extrapulmonary TB. Its high incremental yield and simplicity underscore the need for programmatic integration of LF-LAM with molecular testing to close diagnostic gaps and reduce TB-related mortality.

**Summary Box:** *What is already known about this topic?:*

- Tuberculosis (TB) is the leading cause of death among people living with HIV (PLHIV), contributing to approximately 35% of opportunistic infections among those with advanced HIV disease (AHD).
- Conventional TB diagnosis in PLHIV relies on sputum-based tests such as GeneXpert MTB/RIF, which are often limited by poor sputum production in immunocompromised individuals and reduced sensitivity in extrapulmonary TB.
- The World Health Organization recommends urine-based TB lipoarabinomannan (LF-LAM) testing for PLHIV with AHD, but programmatic evidence from Nigeria remains scarce.

*What this study adds:*

- This study provides further insight on the diagnostic accuracy and yield of Determine Urine TB LF-LAM® among adults and children with AHD in Nigeria in a real programmatic setting.
- It demonstrates the added diagnostic value of urine-based LF-LAM testing in immunocompromised patients, including those unable to produce sputum or with extrapulmonary TB.

*How this study might affect research, practice or policy:*

- Findings support integrating LF-LAM into the routine AHD care package in Nigeria and similar high-burden, resource-limited settings.
- Evidence from this study can inform national HIV and TB program guidelines and accelerate scale-up of non-invasive TB diagnostic strategies to reduce TB-related mortality.

## Introduction

Tuberculosis (TB) is the leading cause of death from a single infectious agent and the most common opportunistic infection among PLHIV [1, 2]. TB substantially accelerates the progression of HIV infection to AIDS, and HIV, in turn, is the primary driver of the TB epidemic in many high-burden countries [1, 3]. It accounts for about 35% of opportunistic infections in individuals with advanced HIV disease (AHD) [4]. In 2023, an estimated 1.25 million people died of TB globally, and approximately 161,000 of these deaths were attributable to TB-HIV coinfection [1].

The burden of TB-HIV coinfection is concentrated in sub-Saharan Africa, which accounted for 73% of all TB-HIV cases and 81% of TB-HIV-related deaths [5]. Only 56% of TB patients living with HIV in 2023 received antiretroviral therapy (ART) [1]. Nigeria, classified among the 30 high-burden countries for TB and TB/HIV, has a particularly severe situation [6]. In 2019, Nigeria reported approximately 46,000 incident TB cases among PLHIV; however, only 12,052 were notified, meaning that nearly three-quarters of cases went undiagnosed and untreated [7, 8]. Such diagnostic gaps represent a major obstacle to achieving both the “End TB Strategy” and the UNAIDS 95-95-95 targets.

Diagnosis remains the weakest link in the TB care cascade, especially among PLHIV with AHD, who are at greatest risk for TB-related morbidity and mortality [9]. In 2022, 30% of TB cases were not diagnosed in Africa region [10] that implies an estimate of 700 000 people missed diagnosis in 2022 alone. AHD is characterized by severe immunosuppression CD4 count below 200 cells/mm^3^, WHO clinical stage 3 or 4 disease, or specific conditions such as cryptococcal meningitis or severe bacterial infections [11]. Individuals with AHD frequently present with nonspecific or atypical TB symptoms, increasing the risk of missed or delayed diagnosis. Moreover, they often have difficulty producing sputum, limiting the effectiveness of conventional diagnostic methods such as smear microscopy or nucleic acid amplification tests [11]. GeneXpert MTB/RIF, the WHO-recommended test for TB diagnosis, relies primarily on sputum samples and is optimized for pulmonary TB detection. Its performance is significantly reduced in extrapulmonary TB, which is more common in immunocompromised individuals, including those with AHD [12]. These limitations underscore the need for complementary diagnostic tools that are easy to use, non-invasive, and effective in detecting TB in severely immunosuppressed patients.

Urine-based TB lipoarabinomannan (LF-LAM) assays represent a promising solution to these challenges. Lipoarabinomannan, a glycolipid component of the Mycobacterium tuberculosis cell wall, is released into the bloodstream and filtered into urine in higher concentrations as TB disease progresses, especially in immuno-compromised patients with advanced HIV infection [13]. Urine testing offers distinct advantages: collection is non-invasive, feasible even in very ill patients or young children, and does not require specialized facilities or highly trained personnel. These characteristics make LF-LAM particularly suitable for use in resource-limited settings and for patients unable to produce sputum [14].

The World Health Organization currently recommends LF-LAM testing for TB diagnosis among PLHIV with signs and symptoms of TB or those with AHD, especially in inpatient or outpatient settings where rapid diagnosis can be lifesaving [15]. Despite these guidelines, evidence on its diagnostic performance under routine programmatic conditions remains limited in many high-burden countries, including Nigeria [16]. Such evidence is essential for informing national policies and ensuring appropriate integration of LF-LAM into the AHD care package to reduce TB-related morbidity and mortality.

This study aims to evaluate the diagnostic accuracy and yield of Determine Urine TB LF-LAM® compared to GeneXpert MTB/RIF among adults and children (<5 years) with AHD enrolled in HIV treatment programs in Nigeria. By providing real-world evidence from programmatic settings, this study seeks to support policy decisions on the role of LF-LAM in strengthening TB diagnosis and care for PLHIV in resource-constrained, high-burden contexts.

## Methods

### Study Design and Setting

We conducted a cross-sectional diagnostic accuracy study across 334 APIN Public Health Initiatives–supported HIV treatment facilities in Nigeria between July 2022 to July 2025. The study followed the Nigeria National HIV and TB guidelines, which recommend LF-LAM testing for all individuals with advanced HIV disease (AHD) and sputum-based GeneXpert MTB/RIF testing for all LF-LAM positives to rule out rifampicin resistance and all presumptive with a negative LF-LAM result. Figure 1

**Figure 1:**
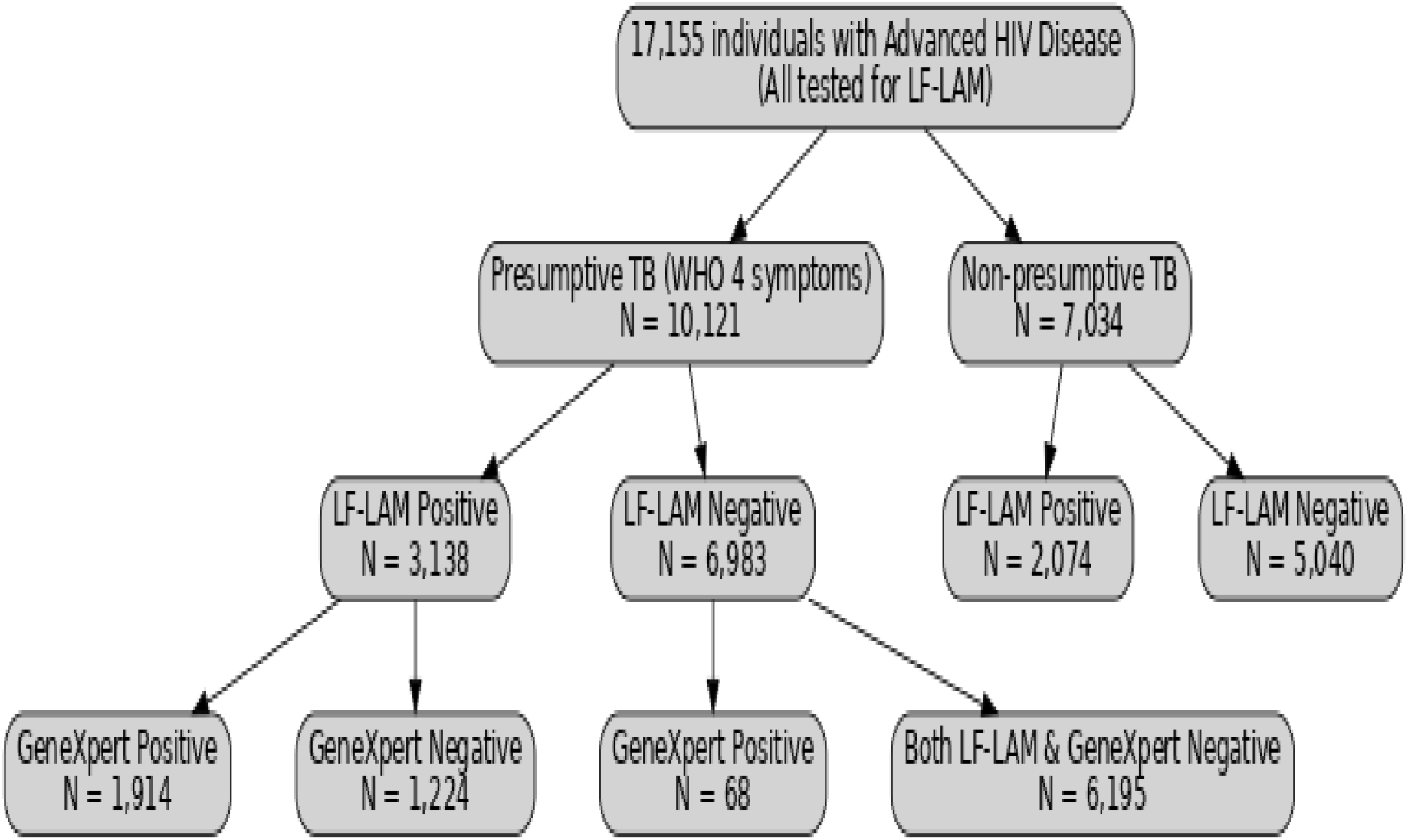
Study sample collection and testing flow

### Study Population

Participants were adults and children living with HIV who met AHD criteria, defined as a CD4 count <200 cells/mm^3^, WHO clinical stage 3 or 4 disease, or serious opportunistic infection or children <5years of age. Individuals were included if they were tested for TB using LF-LAM as part of routine care. Patients with incomplete LF-LAM results or without AHD classification were excluded.

### Sample Size and Sampling

The minimum sample size was estimated using Buderer’s [17] formula for diagnostic accuracy studies. Based on the sensitivity of 60% for LF-LAM, TB prevalence of 35% among AHD patients, 95% confidence, and ±5% precision, the calculated minimum sample size was 1869.

### Procedures

#### Index Test (LF-LAM)

Urine samples were collected and tested using the Determine TB-LAM® Ag lateral flow assay (Abbott Diagnostics, formerly Alere) according to the manufacturer’s instructions. Briefly, 60 μL of unprocessed urine was applied to the sample pad. After 25 minutes, strips were read by two independent trained staff, blinded to patient clinical status and other test results, using the reference card. Discrepancies were resolved by consensus. Test results were scored as grade 1–5 using the reference card. Positive results were defined as grade ≥2 band intensity or negative (grade <2) [18]. Figure 2.

**Figure 2:**
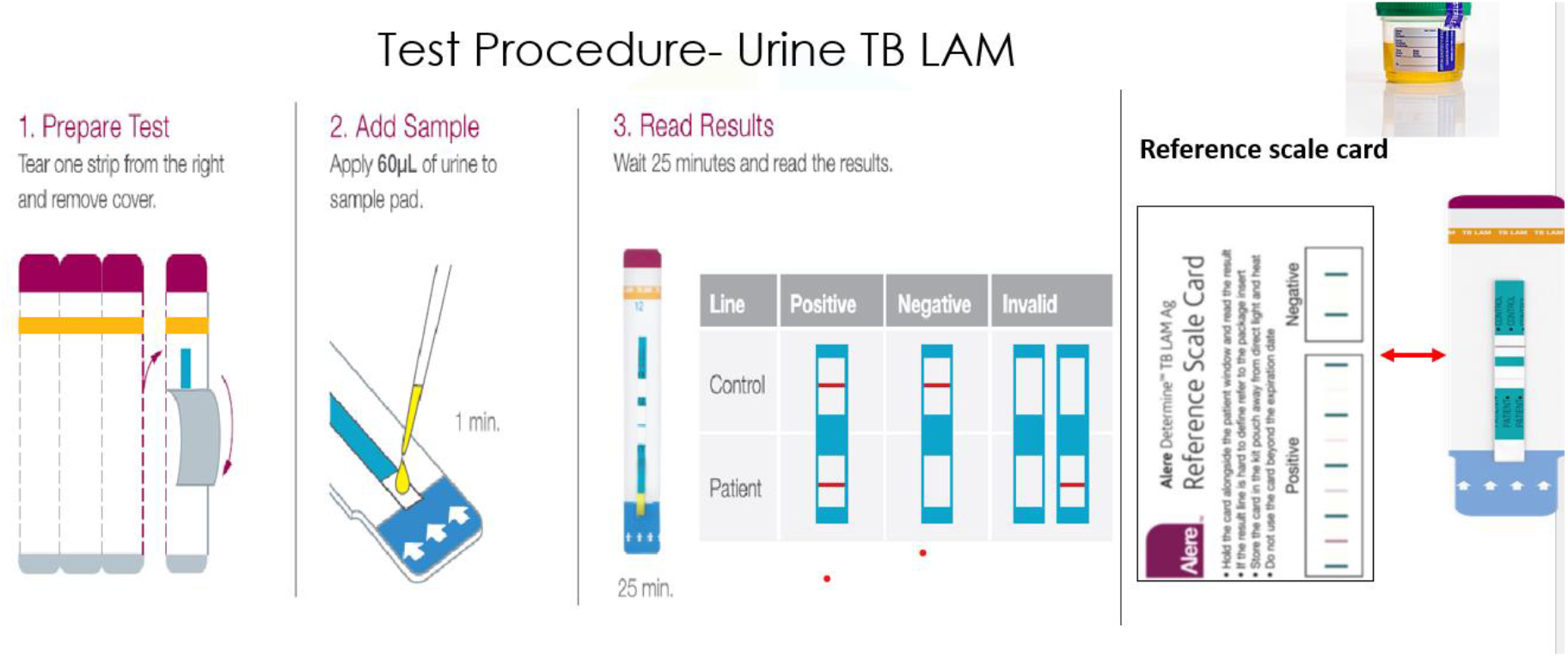
Urine TB LF LAM testing procedures

#### Reference Standard (GeneXpert MTB/RIF)

The primary reference standard was GeneXpert MTB/RIF assay performed on sputum collected from all LF-LAM–positive patients who were presumptive TB cases (based on WHO four-symptom screening). GeneXpert positivity indicated microbiological confirmation of TB [19] figure 3 For sensitivity analysis, a composite reference standard was applied, defined as either GeneXpert positive or LF-LAM positive with initiation of TB treatment within 7 days.

**Figure 3:**
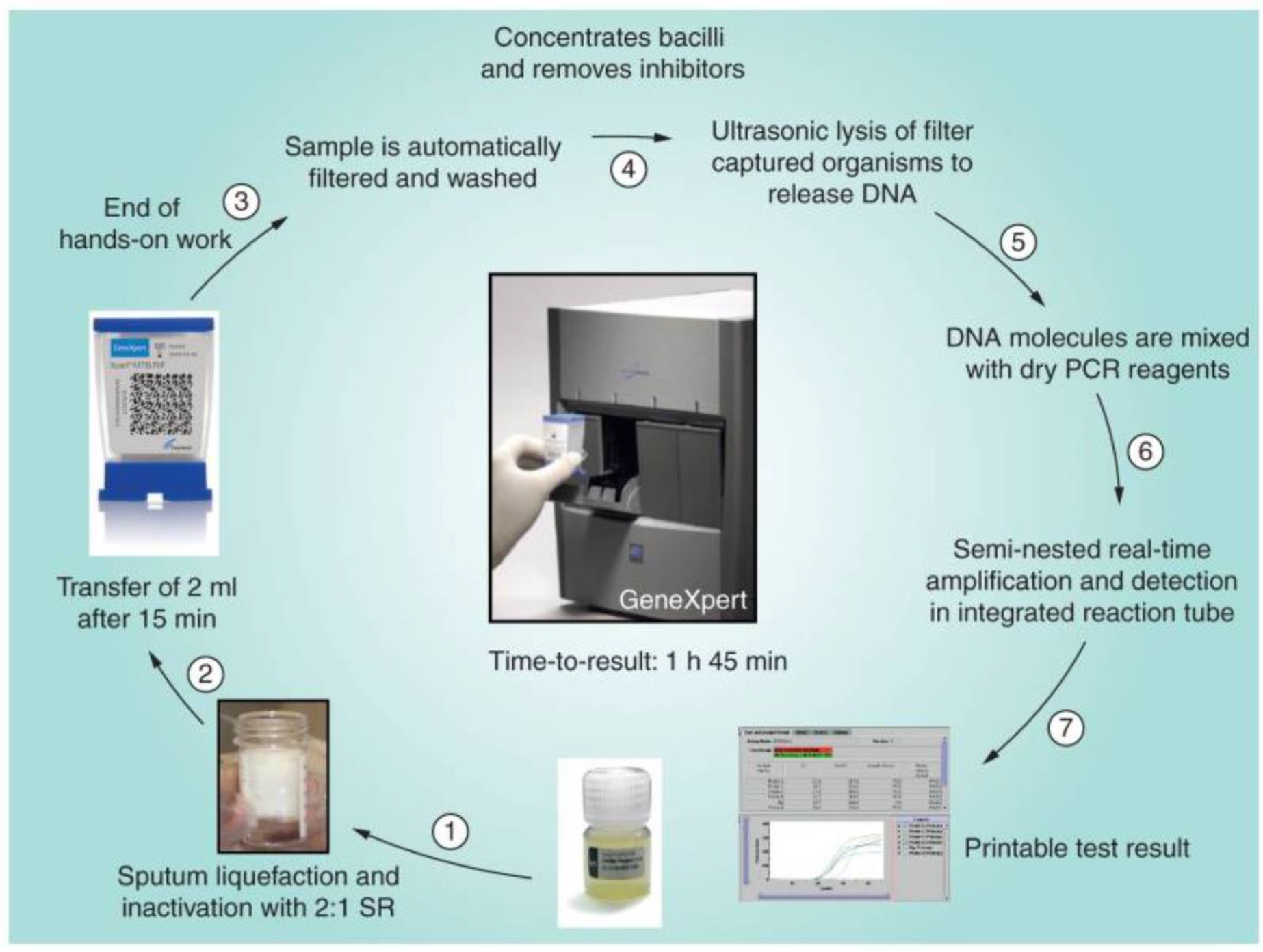
MTB/RIF GeneXpert Testing pr

#### Diagnostic Algorithm and Clinical Management

Following national guidelines, all LF-LAM positive patients were immediately initiated on drug-susceptible TB (DS-TB) treatment while awaiting GeneXpert results. If GeneXpert detected rifampicin resistance, patients would be switched to DR-TB treatment. Patients with GeneXpert-negative results continued DS-TB treatment based on LF-LAM positivity.

#### Data Collection and Management

De-identified programmatic data were extracted from electronic medical records using a structured abstraction tool. Variables included demographics, CD4 count, WHO clinical stage, LF-LAM and GeneXpert results, ART status, and TB treatment initiation.

Data were securely stored on password-protected APIN servers, accessible only to authorized investigators. All identifiers were removed prior to analysis to ensure confidentiality.

#### Statistical Analysis

Diagnostic performance metrics including sensitivity, specificity, positive predictive value (PPV), negative predictive value (NPV), and overall accuracy were computed with 95% confidence intervals (CIs) using the Wilson score method. Agreement between LF-LAM and GeneXpert was assessed using Cohen’s kappa statistic with 95% CIs. Differences in paired classifications between LF-LAM and GeneXpert among individuals tested with both methods were evaluated using McNemar’s chi-squared test.

Diagnostic yield (DY) was defined as the proportion of TB cases identified by a single diagnostic test on the first diagnostic sample collected.

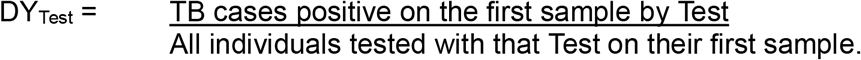

Incremental diagnostic yield (IDY) in paired testing was defined as the proportion of TB cases positive by LF-LAM but negative by GeneXpert divided by all TB cases detected by either test on first samples

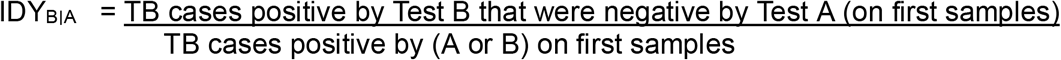

Proportions were presented with 95% CIs using the Wilson score method. A secondary analysis using a composite reference standard (either GeneXpert positive or LF-LAM positive with initiation of TB treatment within 7 days) was performed to evaluate robustness of sensitivity estimates. All analysis was conducted using SPSS version 27 Chicago IL USA.

#### Reporting Guidelines

This study adhered to the STARD (Standards for Reporting Diagnostic Accuracy Studies) guidelines for diagnostic accuracy research [20]

#### Ethical Considerations

Participant demographic and clinical data were recorded using standardized forms and entered in a secure, password-protected database. Diagnostic performance metrics were calculated using standard formulas, with 95% confidence intervals. The study was approved by National Health Ethic Committee (NHREC/01/01/2007-08/05/2025iiB) and by Institutional Review Board of APIN Public Health Initiatives NHREC/APIN-HREC/2/12/24iii.

## Results

### Baseline characteristics of study participant

A total of 17,155 participants with advanced HIV disease (AHD) were tested using LF-LAM across APIN-supported facilities. 5212 (30.4%) were positive for TB. The median age was 36 years (IQR: 28–48), and 61% were female (n=10,465). Children aged <5 years accounted for 1.3% (n=231), and those aged 5–10 years comprised 10% (n=1,716), 91% (n = 15,611) of the participants are ART-naive. WHO clinical staging showed that 12.3% were Stage 3 and 8.7% were Stage 4. Among all participants, 38.4% (n=6594) were classified as presumptive TB by WHO four-symptom screening, with cough (41%), fever (71%), weight loss (57%) and night sweat (31%) being the most frequent symptoms. Table 1.

### Diagnostic Accuracy of LF-LAM vs GeneXpert

The overall diagnostic yield (DY) of LF-LAM on the first urine sample among all AHD patients was 30.4% (5,212/17,155; 95% CI: 28.1–32.8). Notably, 2,074 (40%) of LF-LAM positives were not presumptive by WHO four-symptom screening and would have been missed if symptom screening alone was used prior to GeneXpert testing.

In the paired-tested cohort of 3,138 presumptive TB patients who were LF-LAM positive and had sputum collected for GeneXpert, 1,914 (61.0%) were GeneXpert-positive, and 1224 (39.0%) were GeneXpert-negative but LF-LAM-positive on first samples. The incremental diagnostic yield of LF-LAM over GeneXpert was 39% (1224/ [1,914+1224]; 95% CI: 37.6 –41.4), with significant discordance between the two methods (McNemar’s p<0.001). Table 2.

### Incremental Yield of LF-LAM in TB detection beyond WHO symptoms screenings

Of the 5,212 confirmed TB cases, LF-LAM detected 2,074 TB cases (40% of LF-LAM positives) that would have been missed using WHO symptom-based screening alone, as these individuals were not presumptive by WHO criteria. Additionally, among presumptive TB cases, LF-LAM identified 896 TB cases beyond those confirmed by GeneXpert, contributing to diagnostic yield of LF-LAM over GeneXpert of 39%. The addition of urine LF-LAM to sputum GeneXpert increased the diagnostic yield from 11.6% to 30.8%, an absolute gain of 19.2% (*n* = 3,298/17,155: a 1.6-fold increase; *P* < 0.001). Table 3.

### Stratified Diagnostic Performance of LF LAM

When performance was evaluated across CD4 strata of the reference standard LF-LAM sensitivity was highest in patients with in severely immunocompromised and in children under 5 years of age. Specificity remained high across the strata however it decreased to less than 90% in treatment naïve participants. Table 4.

## Discussion

### Principal Findings

In this large, programmatic evaluation among over 17,000 individuals with advanced HIV disease (AHD) in Nigeria, the Determine TB-LAM assay demonstrated high diagnostic yield and acceptable diagnostic accuracy when benchmarked against a composite reference standard. LF-LAM identified 5,212 TB cases, including 2,074 (40%) among patients without WHO-defined presumptive symptoms, who would otherwise have been missed using symptom-based screening alone. Among presumptive cases, LF-LAM concurred with GeneXpert in detecting 1,914 TB cases, while capturing an additional 1,224 cases not confirmed by GeneXpert but initiated on TB treatment. Overall, the addition of urine LF-LAM to sputum GeneXpert increased the diagnostic yield from 11.6% to 30.8%, representing an absolute gain of 19.2% (3,298 additional TB cases detected). The assay achieved a sensitivity of 76–92% across strata and a specificity consistently above 90%, with particularly high sensitivity among children and those with CD4 <100 cells/mm^3^. These findings reinforce LF-LAM’s role as a rapid, point-of-care tool to close diagnostic gaps in AHD populations. Incremental yield analysis demonstrated that LF-LAM added substantial value beyond symptom screening and GeneXpert, particularly in high-burden, low-resource settings. The sensitivity and specificity estimates should be interpreted with caution. In our programmatic workflow sputum GeneXpert testing was not applied uniformly: GeneXpert was preferentially performed on patients who were LF-LAM positive (and on presumptive symptomatic patients who are LF LAM positive), so verification was conditional rather than independent. This selective verification, together with differential sample sizes and missing data across strata, may have been reason for overestimated sensitivity of urine LF LAM and underestimated the specificity rather than a true LF-LAM sensitivity/specificity over GeneXpert. For these reasons we emphasise the diagnostic yield as the primary programmatic measures.

### Comparison with Other Studies

Our findings are consistent with and expand on prior evidence from diverse high-burden settings. In South Africa, Lawan *et al*. reported that LF-LAM identifies between 20–35% additional TB cases missed by conventional diagnostic pathways [21], aligning with the 39% yield increase observed in our cohort. The study also reported addition of urine LF-LAM to sputum GeneXpert increased diagnostic yield from 26.6% to 52.5%, closely mirroring the 11.6% to 30.8% a 1.6-fold increase p < 0.001 and 19.2% absolute gain observed in our study. Songkhla *et al*. showed LF-LAM specificity of 76.0%, with markedly higher sensitivity 90.5% and NPV 96.4% among patients with CD4 <50/µL [22], findings that align with our composite analysis in which sensitivity was 90.0% among those with CD4 <100 cells/mm^3^ while specificity remained high across strata.

Similar to our study, Nathavitharana *et al*. reported improved yield in hospitalized HIV-positive patients, particularly those with low CD4 counts [23]. However, our study is unique as it reflects real-world programmatic implementation across multiple facilities, rather than controlled clinical trials. Unlike prior studies, we quantified the incremental yield in non-presumptive patients, highlighting LF-LAM’s value in routine care beyond inpatient settings.

### Discordance between Urine LF-LAM and GeneXpert

A key finding of this study is the discordance observed between LF-LAM and GeneXpert results. Of the 3,138 LF-LAM positive presumptive cases, 1,224 (39%) were GeneXpert negative, while 68 cases were presumptive to TB, LF-LAM negative but GeneXpert positive. This discordance is not unexpected and reflects the biological and operational differences between the two assays. LF-LAM detects mycobacterial lipoarabinomannan antigen excreted in urine, which is particularly elevated in individuals with extrapulmonary TB and profound immunosuppression. In contrast, GeneXpert is primarily a sputum-based assay, in our setting optimised for pulmonary TB, and may miss cases where bacillary burden is low in sputum, but disease is disseminated. This explains why many LF-LAM positive, but GeneXpert negative patients were nonetheless initiated on TB treatment, consistent with WHO recommendations for LF-LAM use in AHD populations. The 68 GeneXpert positive but LF-LAM negative cases likely represent individuals with pulmonary TB but lower antigenuria, again illustrating complementary rather than redundant diagnostic profiles of the two assays. Similar discordance has been reported in studies from Uganda, South Africa, and Asia [21,22,23], where LF-LAM consistently identified TB in severely immunocompromised patients missed by sputum-based diagnostics, while GeneXpert captured some pulmonary cases not detected by LF-LAM. Clinically, this discordance underscores the importance of using LF-LAM alongside GeneXpert rather than as a replacement. The combined approach maximises diagnostic yield. In our study, addition of LF-LAM to routine GeneXpert increased case detection from 11.6% to 30.8%. This supports WHO’s updated guidelines recommending LF-LAM as a complementary tool in AHD diagnostic algorithms.

### Clinical, programmatic, and policy implications

Our findings have several important clinical and programmatic implications. First, the detection of 2,074 TB cases among non-presumptive AHD patients demonstrates that reliance on WHO symptom screening alone misses a substantial proportion of disease. Incorporating LF-LAM into routine care allows clinicians to diagnose TB earlier, including extrapulmonary disease, and initiate timely treatment without waiting for sputum results, thereby reducing morbidity and mortality in a highly vulnerable population.

Second, the observed discordance between LF-LAM and GeneXpert highlights the complementary nature of the two assays. LF-LAM provided additional diagnostic value for disseminated TB in immunosuppressed patients, while GeneXpert retained value for pulmonary TB cases with low antigenuria. From a clinical perspective, our findings highlight the need to integrate LF-LAM alongside GeneXpert in routine AHD care, since each test identifies cases the other misses. A combined approach maximises diagnostic yield while ensuring detection of both pulmonary and extrapulmonary TB.

Programmatically, our evaluation demonstrates the feasibility of scaling up LF-LAM across a national HIV program. Testing was integrated into routine workflows without major disruption, and results were available at the point of care, supporting rapid treatment initiation. The incremental diagnostic yield of 19.2% translates into thousands of additional TB cases detected and treated, which has the potential to significantly improve TB case finding among HIV co-infected and reduce TB-related mortality among PLHIV. Furthermore, by identifying TB earlier, LF-LAM may reduce downstream costs associated with advanced disease management, prolonged hospitalisation, and transmission.

At the policy level, these findings provide real-world evidence in support of Nigeria’s decision to include LF-LAM testing in its National HIV and TB guidelines. They also offer broader lessons for other high-burden countries considering LF-LAM scale-up. Given the WHO recommendation that LF-LAM be used for all PLHIV with AHD or CD4 <200 cells/mm^3^, our data strongly support prioritising procurement, training, and implementation at scale. Moreover, aligning LF-LAM deployment with Global Fund and PEPFAR programmatic support could accelerate progress towards global End TB targets.

### Strengths and Limitations

Strengths of this study include its large sample size, real-world programmatic setting integrating with point-of-care CD4 and other diagnostic tools, and systematic application of WHO-recommended testing protocols. We adhered to STARD guidelines and performed stratified analyses by CD4 count, ART status, and age. However, limitations exist. Specificity was underestimated due to conditional sputum testing only for LF-LAM positive cases, introducing verification bias. Additionally, use of a composite reference standard may overestimate sensitivity. Finally, the study did not assess patient outcomes post-diagnosis, which would provide further insight into clinical impact and mortality.

### Future Research

Future studies should assess the impact of LF-LAM on patient outcomes, including survival and treatment success, in real-world programs. Evaluating cost-effectiveness and implementation barriers will further inform scale-up across Nigeria and sub-Saharan Africa. Importantly, studies integrating LF-LAM with novel non-sputum diagnostics, including urine-based molecular assays, could optimise AHD diagnostic algorithms. Longitudinal research linking LF-LAM results with clinical outcomes will also strengthen the evidence base for routine use beyond current WHO target groups.

## Conclusion

This study confirms that LF-LAM adds diagnostic value to GeneXpert in routine HIV care in Nigeria, particularly for severely immunocompromised and hospitalized patients. While LF-LAM cannot replace GeneXpert due to its lower sensitivity, its adjunctive use enhances TB case detection and supports earlier treatment initiation, aligning with global TB/HIV collaborative strategies.

## Data Availability

All data produced in the present study are available upon reasonable request to the authors

https://apinnigeria-my.sharepoint.com/:x:/g/personal/jnwofe_apin_org_ng/EY0Y5L2zZSRLsB26v8k1FUoBNT97euUlk0Wu_rshkzbLNQ?e=R6o4sU

## Acknowledgement

We sincerely thank the management and staff of the participating APIN-supported health facilities in Benue, Plateau, Ogun, Oyo, and Ondo States for their invaluable support during this study. We acknowledge the dedication of all laboratory scientists, clinicians, and other healthcare workers who participated involved in the study. We are grateful to the patients and their caregivers for their willingness to participate in this study.

## Funding

The authors received no funding for this study

## Conflict of Interest

The authors declare that they have no competing interests

## Authors’ contributions

JON conceived the study. Protocol development was undertaken by JON, DCO, DAA, EAO, and FEO. Data collection and laboratory testing were performed by DO, PCO, IOO, DCO, AMO, OSO, OO, AIC, JOO, MO, AIC, and AA. Data analysis was conducted by DAA, EE, II, JON, DCO and LIM. The manuscript was drafted by JON, COU, DCO, DAA, II, LIM, FEO, OK, LIM, DSA, II, and DCO. All authors contributed to the review of the final version of the manuscript. JON, FEO, EAO, DCO, COU, II, DAA, and EO reviewed final copy for submission. All authors agree to be accountable for all aspects of the work.

## Notes

### Competing Interest Statement

The authors have declared no competing interest.

### Funding Statement

This study did not receive any funding

### Author Declarations

The study was approved by the National Health Ethic Committee of Nigeria(NHREC/01/01/2007-08/05/2025iiB)

