## Supplemental Tables for "Diagnostic Accuracy and Yield of Determine Urine TB LF-LAM® for Detecting Tuberculosis Among Adults and Children with Advanced HIV Disease in Nigeria"

**Table 1: Baseline characteristics of study participants**

| Characteristic | All (n=17,155) | Presumptive TB (n=6594) | Confirmed TB (n=5,084) | Non-TB (n=12,097) |
|---|---|---|---|---|
| **Sex** | | | | |
| Male | 6,690 (39.0%) | 2,572 (39.0%) | 1,973 (38.8%) | 4,717 (39.0%) |
| Female | 10,465 (61.0%) | 4022 (61.0%) | 3,085 (61.2%) | 7,380 (60.0%) |
| **Age** | | | | |
| *Median (IQR), years* | 36 (28–48) | – | – | – |
| <5 years | 231 (1.3%) | 46 (0.7%) | 81 (1.6%) | 150 (1.2%) |
| 5–10 years | 1,716 (10.0%) | 659 (10.0%) | 485 (9.5%) | 1,190 (9.8%) |
| 10–20 years | 2,685 (15.7%) | 1055 (16.0%) | 799 (15.7%) | 1,926 (15.9%) |
| 30–40 years | 5,891 (34.3%) | 2242 (34.0%) | 1,720 (33.8%) | 4,113 (34.0%) |
| >50 years | 6,632 (38.7%) | 2591 (39.3%) | 1,973 (38.8%) | 4,718 (39.0%) |
| **CD4 (cells/mm³)** | | | | |
| *Median (IQR)* | 64 (4–200) | – | – | – |
| <50 | 665 (3.9%) | 151 (2.3%) | 313 (6.2%) | 352 (2.9%) |
| 50–100 | 1,414 (8.2%) | 1266 (19.2%) | 609 (12.0%) | 805 (6.7%) |
| 100–200 | 2,079 (12.1%) | 1800 (27.3%) | 921 (18.1%) | 1,158 (9.6%) |
| ≤200† | 10,878 (63.4%) | 3,376 (51.2%) | 2,234 (43.9%) | 8,644 (71.5%) |
| **WHO clinical stage** | | | | |

| Characteristic | All (n=17,155) | Presumptive TB (n=6594) | Confirmed TB (n=5,084) | Non-TB (n=12,097) |
|---|---|---|---|---|
| Evaluated for WHO stagging | 8234 (48.0%) | | | |
| Stage 3 | 1,013 (12.3%) | 234 (3.7%) | 519 (10.2%) | 494 (16.3%) |
| Stage 4 | 719 (8.7%) | 171 (2.6%) | 462 (9.1%) | 257 (2.1%) |
| **TB symptom screening (WHO)** | | | | |
| Screened for TB symptoms | 15,954 / 17,155 (93.0%) | – | – | – |
| Cough‡ | 6,541 (41.0%) | 1,432 (22.0%) | 912 (18.0%) | 5,629 (46.0%) |
| Fever‡ | 11,327 (71.0%) | 2,417 (36.7%) | 570 (11.2%) | 10,757 (89.0%) |
| Weight loss‡ | 9,094 (57.0%) | 1,820 (27.6%) | 1,112 (22.0%) | 7,982 (66.0%) |
| Night sweats‡ | 4,946 (31.0%) | 925 (14.0%) | 390 (7.7%) | 4,556 (38.0%) |
| Previous TB history‡ | 479 (3.0%) | 0 | 0 | 0 |
| **ART Status** | | | | |
| ART Naïve | 15,611 (91.0%) | 2383 (80.0%) | 4595 (90.4%) | 11,016 (91.1%) |
| ART Experience | 1,544 (9.0%) | 601 (20.0%) | 489 (9.6%) | 1081 (8.9%) |

Note!. Screened for presumptive is higher than “n” when you add up all the individuals with various symptom because an individual can present with more than one of the WHO symptoms for TB.

**Table 2. Diagnostic accuracy of LF-LAM vs GeneXpert**

| **LF-LAM** | **GeneXpert +** | **GeneXpert −** | **Total** |
|---|---|---|---|
| Positive | 1,914 | 1,224 | 3,138 |
| Negative | 68 | 6,195 | 6,263 |
| **Total** | 1,982 | 7,419 | 10,121 |

| **Metric** | **Estimate** | **95% CI** |
|---|---|---|
| Sensitivity | 96.6% | 95.8–97.2 |
| Specificity | 83.5% | 82.6–84.4 |
| PPV | 61.0% | 59.4 –62.6 |
| NPV | 98.9% | 98.6–99.2 |
| Overall Accuracy | 85.0% | 84.3–85.8 |
| Cohen’s Kappa | 0.66 | — |
| McNemar p-value | <0.001 | — |

**Table 3: Incremental Yield of LF-LAM in TB detection beyond WHO symptoms screenings**

| Measure | Value | 95% CI |
|---|---|---|
| LF-LAM DY (all AHD patients) | 30.4% (5,212/17,155) | 28.1–32.8 |
| LF-LAM positives non-presumptive | 40.0% (2,074/5,212) | — |
| GeneXpert positives (paired cohort) | 61.0% (1,914/3,138) | — |
| LF-LAM positives, GeneXpert negative | 39.0% (1224/3,138) | — |
| Incremental diagnostic yield of LF-LAM | 39% (1224/ [1,914+1224]) | 37.6–41.4 |

**Table 4: Stratified Diagnostic Accuracy of LF-LAM using a Composite Reference standard**

| **Stratum** | **Sensitivity % (95% CI)** | **Specificity % (95% CI)** |
|---|---|---|
| **CD4 <100** | 81.8 (78.8–84.6) | 100 (98.0–100) |
| **CD4 100–199** | 93.8 (92.5–95.0) | 91.2 (89.0–93.0) |
| **ART-naïve** | 89.1 (87.3–90.7) | 88.4 (84.9–91.2) |
| **ART-experienced** | 91.2 (88.6–93.3) | 96.2 (94.2–97.6) |

| Stratum | Sensitivity % (95% CI) | Specificity % (95% CI) |
|---|---|---|
| **Age <5 years** | 96.4 (89.7–98.9) | 100 (84.5 – 100) |
| **Age ≥5 years** | 89.6 (88.2–91.0) | 92.8 (91.0–94.3) |
